# A Network Medicine Approach to Investigating ME/CFS Pathogenesis in Severely Ill Patients: A Pilot Study

**DOI:** 10.1101/2024.09.26.24314417

**Authors:** Li-Yuan Hung, Chan-Shuo Wu, Chia-Jung Chang, Peng Li, Kimberly Hicks, Becky Taurog, Joshua J Dibble, Braxton Morrison, Chimere L Smith, Ronald W Davis, Wenzhong Xiao

## Abstract

This pilot study harnessed the power of network medicine to unravel the complex pathogenesis of Myalgic Encephalomyelitis/Chronic Fatigue Syndrome (ME/CFS). By utilizing a network analysis on whole genome sequencing (WGS) data from the Severely Ill Patient Study (SIPS), we identified ME/CFS-associated proteins and delineated the corresponding network-level module, termed the SIPS disease module, together with its relevant pathways. This module demonstrated significant overlap with genes implicated in fatigue, cognitive disorders, and neurodegenerative diseases. Our pathway analysis revealed potential associations between ME/CFS and conditions such as COVID-19, Epstein-Barr virus (EBV) infection, neurodegenerative diseases, and pathways involved in cortisol synthesis and secretion, supporting the hypothesis that ME/CFS is a neuroimmune disorder. Additionally, our findings underscore a potential link between ME/CFS and estrogen signaling pathways, which may elucidate the higher prevalence of ME/CFS in females. These findings provide insights into the pathogenesis of ME/CFS from a network medicine perspective and highlight potential therapeutic targets. Further research is needed to validate these findings and explore their implications for improving diagnosis and treatment.

## 1 Introduction

Myalgic Encephalomyelitis/Chronic Fatigue Syndrome (ME/CFS) is a complex and debilitating condition characterized by persistent fatigue, cognitive impairments, post-exertional malaise, and a variety of other symptoms that severely impact patients’ quality of life (Cortes Rivera et al., 2019). Notably, the condition predominantly affects women (Lim et al., 2020). Despite extensive research, the etiology of ME/CFS remains elusive, and effective treatments are still lacking. Recent evidence suggests that neuroimmune dysfunction plays a crucial role in the pathogenesis of ME/CFS (Marshall-Gradisnik and Eaton-Fitch, 2022; Morris and Maes, 2013a, 2013b; Ryabkova et al., 2022).

Genetic factors are believed to contribute significantly to ME/CFS, with prior studies suggesting a genetic predisposition to the disease (Afari and Buchwald, 2003; Albright et al., 2011). However, the specific genes and pathways involved have not been fully elucidated (Dibble et al., 2020). This study employs a network medicine approach to investigate the genetic underpinnings of ME/CFS, focusing on its most severe manifestations.

Approximately 25% of ME/CFS patients are severely ill and are often housebound or bedridden (Feiden, 1990; Fennell et al., 1998; Pendergrast et al., 2016). Our research centers on the Severely Ill Patient Study (SIPS) cohort, which includes patients enduring the most severe symptoms of ME/CFS (Chang et al., 2021). These individuals suffer from profound fatigue, cognitive impairment, and a significantly reduced quality of life. Previous analyses of this cohort revealed unique sleep profiles, cognitive deficits, and markedly low morning cortisol levels. Additionally, parallels between ME/CFS and long COVID symptoms suggest common etiological pathways (Chang et al., 2021).

The credibility of the SIPS cohort is reinforced by its validation by medical professionals, ensuring the authenticity and representativeness of the data. The network medicine approach provides a sophisticated framework for understanding disease mechanisms at the molecular level. This paradigm introduces the concept of *disease modules*, interconnected subgraphs within the protein-protein interaction (PPI) network that represent the molecular facets of specific conditions. By examining these modules, researchers can predict disease relationships and elucidate the molecular mechanisms underlying complex diseases (Barabási et al., 2011; Menche et al., 2015; Sharma et al., 2015).

By harnessing the power of PPI network analysis and whole genome sequencing (WGS), we identified pathways potentially contributing to ME/CFS development. Our findings suggest significant involvement of the immune and neurological systems, indicating shared genetic underpinnings with these systems.

In summary, this study sheds light on the genetic landscape of ME/CFS, providing new insights and potentially paving the way for novel therapeutic interventions. Our findings, particularly those related to immune and neurological pathways, could reshape our understanding of ME/CFS and lead to more targeted treatment strategies.

## 2 Results

In the Severely Ill Patient Study (SIPS) (Chang et al., 2021), we investigated the medical conditions of 20 ME/CFS patients exhibiting severe symptoms, as previously documented. These individuals met the International Consensus Criteria for ME/CFS (ME-ICC) (Carruthers et al., 2011) and were confined to their homes, remaining sedentary and reclined for over 14 hours daily. Their SF-36 physical functioning scores and Karnofsky Performance Status Index (Karnofsky and Burchenal, 1949) scores were both below 70.

WGS was performed on the blood samples from the 20 SIPS patients, and high-confidence variants were filtered based on several criteria (see Methods), resulting in 2,798,019 variants for further analysis. Using the American College of Medical Genetics and Genomics (ACMG) criteria, we identified 103 pathogenic or likely pathogenic variants (Qiagen IVA). These were then compared to background population data, leading to the removal of 7 common SNVs. The remaining 96 variants were aggregated at the gene level (Table S1). Among them, 9 genes were found to affect at least 2 (10%) of the 20 patients (Figure 1, Table S1).

**Figure 1.**
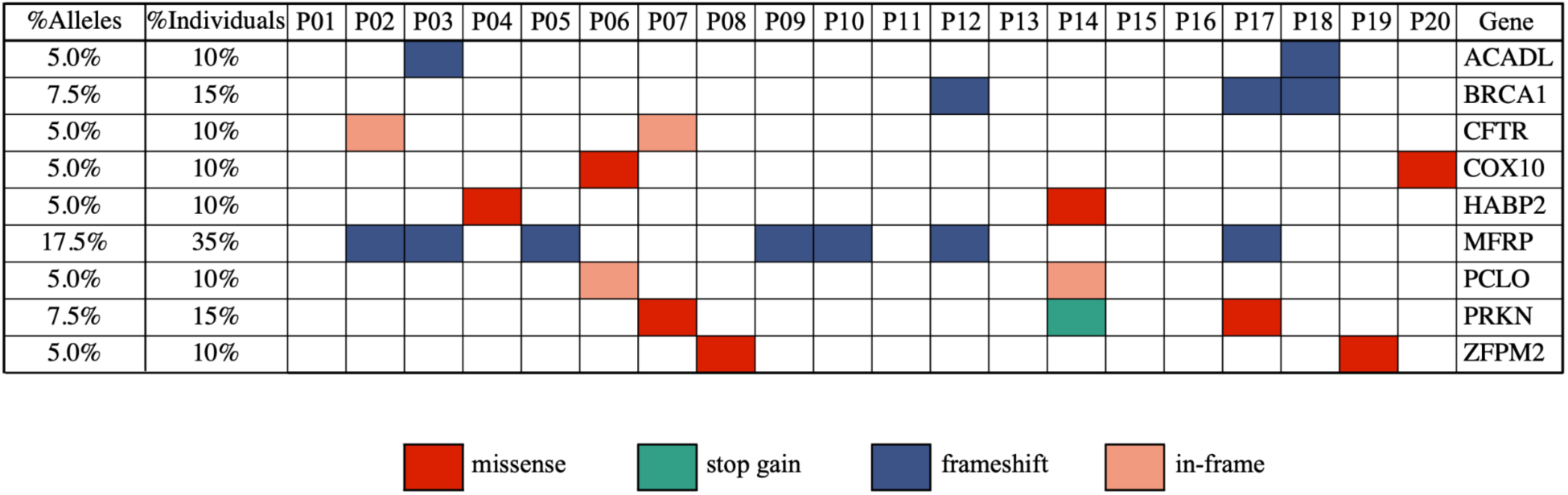
Identification of recurrent pathogenic or likely pathogenic variants (using the ACMG criteria) in 20 severely ill ME/CFS patients.

We examined the nine genes with recurrent variants in the SIPS patients (Figure 1), specifically *ACADL*, *BRCA1*, *CFTR*, *COX10*, *HABP2*, *MFRP*, *PCLO*, *PRKN*, and *ZFPM2*. Given the defining characteristic of cognitive abnormalities in SIPS patients, it was crucial to assess the relevance of these genes to their symptoms. Seven of the nine genes (77.8%) have previously been linked to cognitive impairment or neurological functions. For instance, *BRCA1* is involved in maintaining genomic stability and has implications in neurodegeneration and stress-induced senescence (Leung and Hazrati, 2021; Suberbielle et al., 2015). The *PRKN* gene is associated with Parkinson’s disease, characterized by both motor and non-motor symptoms (Lücking et al., 2000). *CFTR*’s expression in the nervous system may influence mood, memory, energy balance, olfaction, motor function, respiration, and autonomic control of visceral organs (Reznikov, 2017). Mutations in the *COX10* gene, crucial for cellular respiration, have been linked to various peripheral neuropathy disorders (Fünfschilling et al., 2012; Kuroha et al., 2022). *MFRP* is tied to eye development, with mutations affecting retinal microglia (Kumari et al., 2022). *PCLO*, encoding the Piccolo protein essential for synaptic transmission, has mutations linked to pontocerebellar hypoplasia, a rare neurodegenerative disorder (Ackermann et al., 2019; Ahmed et al., 2015). *ZFPM2* is involved in the development of GABAergic neurons in specific brain regions (Morello et al., 2020).

Our results suggest a connection between the identified pathogenic variants and cognitive symptoms in the SIPS patients. While further studies are required to elucidate how these genes influence cognitive impairment in the SIPS patients, Figure 1 provides a valuable foundation for future investigations.

To investigate the genetic factors contributing to the onset and progression of ME/CFS in severely ill patients, we utilized a network-based approach that considers the interconnections between disease-related genes. This approach aims to identify and locate the SIPS disease module within the human PPI network and discern its interplay with other pathways or conditions. During the networ construction process, emphasis is placed on relevant genes, as disease-associated genes tend to cluster together, forming interconnected subnetworks, while unrelated genes scatter randomly and minimally influence the subnetwork’s structure (Menche et al., 2015).

Applying the Disease Module Detection (DIAMOnD) algorithm (Ghiassian et al., 2015), we successfully inferred the SIPS disease module by starting with a subgraph of seed genes derived from 96 pathogenic or likely pathogenic variants identified in the 20 SIPS patients (Table S1). Using this set of seed genes as the input, the DIAMOnD algorithm identified the top 200 neighborhoods, forming the SIPS disease module (for details, refer to Table S2). Within the disease module, we noted a significant presence of genes related to fatigue and cognitive disorders. Using the disease-gene association data from DisGeNET (Piñero et al., 2020) as a reference, genes linked to fatigue exhibited a 3.94-fold enrichment (p-value: 1.3899e-14, cumulative hypergeometric test), while those connected to cognitive disorders showed a 2.29-fold enrichment (p-value: 0.0005) (Figure 2A). Furthermore, the SIPS disease module was found to intertwine with the six neurodegenerative disease pathways in KEGG (Kanehisa et al., 2023) (Figure 2A, 2B), with enrichment ranging from 2.48 to 4.61 fold (Figure 2A).

**Figure 2.**
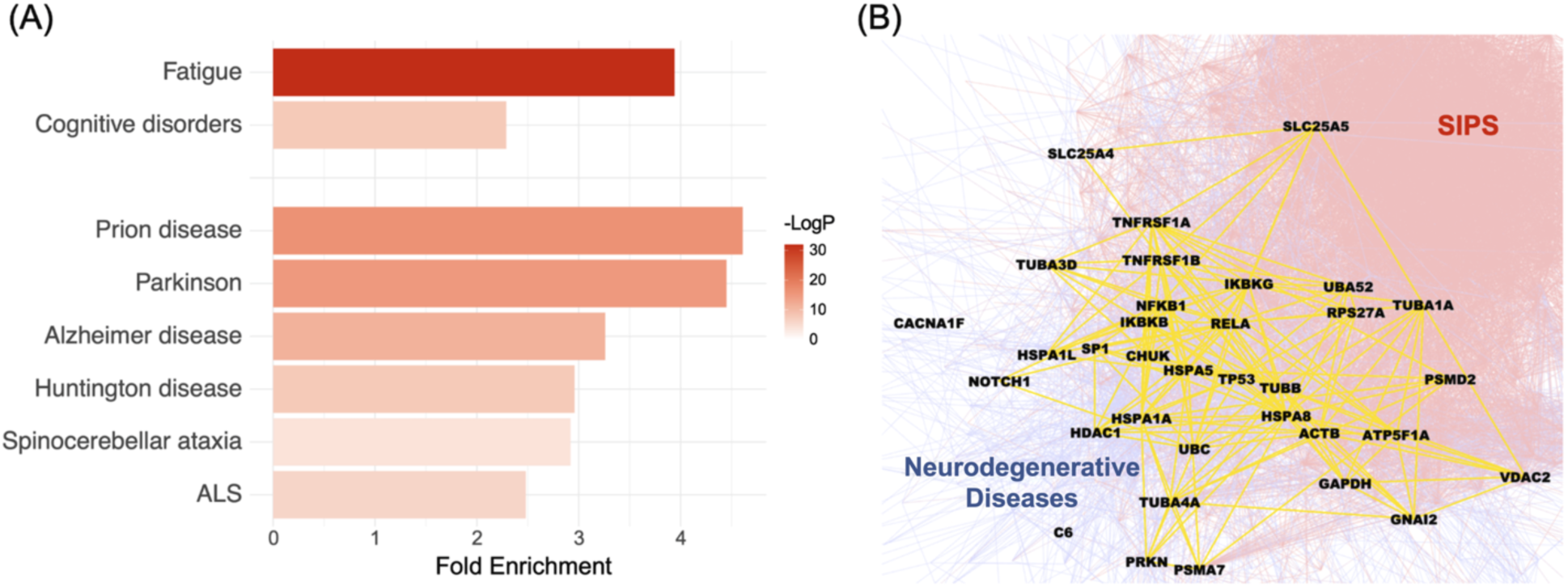
Examining the relationship between the SIPS disease module and ME/CFS diagnostic phenotypes, as well as the underlying biological mechanisms. (A) The significance of the enrichment between the SIPS disease module, two ME/CFS underlying conditions (fatigue and cognitive disorders) and six specific neurodegenerative diseases (Prion disease, Parkinson’s Disease, Alzheimer’s disease, Huntington’s disease, Spinocerebellar ataxia, and ALS). The bar plots show fold enrichments, and the colors show the p-values. (B) The interrelationship between the SIPS disease module and neurodegenerative disease pathways from KEGG. The intersection between these two entities is represented in yellow, with the SIPS side in red and the neurodegenerative disease side in blue.

Considering the widespread cognitive abnormalities in severely ill ME/CFS patients and the unresolved questions surrounding the underlying mechanisms of these conditions, the intersection of the SIPS disease module with neurological disease pathways is especially noteworthy.

In our subsequent analysis, we compared the SIPS disease module with the 70 disease modules presented in the DIAMOnD publication (Ghiassian et al., 2015). These 70 diseases were classified into 4 distinct categories based on their conceivable links to the hypothesized mechanisms of ME/CFS (Marshall-Gradisnik and Eaton-Fitch, 2022). A fifth category, cancer, was incorporated as an out-group for comparative purposes. Our findings revealed that certain diseases exhibited similarities with the SIPS disease module, as depicted in Figure 3A. Notably, nearly half of the top 20 diseases showing the greatest resemblance were either entirely or partially neurological disorders. These included conditions such as muscular dystrophies, basal ganglia diseases, spinocerebellar degenerations, Charcot−Marie−Tooth disease, motor neuron disease, cerebrovascular disorders, amyotrophic lateral sclerosis (ALS), and cerebellar ataxia. In contrast, cancer disease modules showed minimal similarity to the SIPS disease module. When visualizing the SIPS disease module alongside the 70 DIAMOnD disease modules in a network diagram, it was evident that the SIPS module closely overlapped with modules related to immune and neurological systems (Figure 3B). The predominance of immune and neurological conditions among the five most similar diseases suggests that the SIPS disease module might share underlying causes with these two systems.

**Figure 3.**
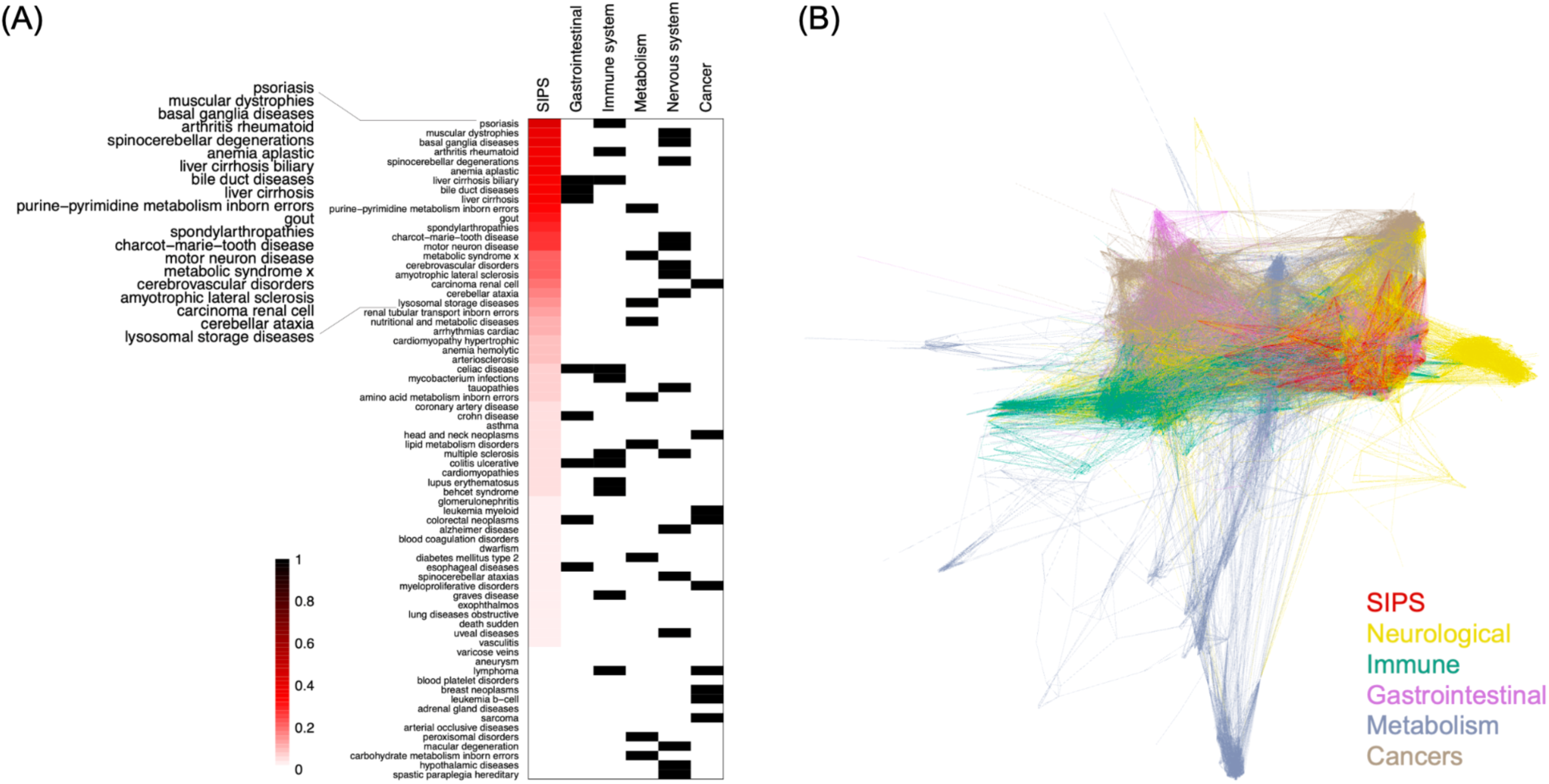
The SIPS and DIAMOnD disease modules. (A) The relationship between the SIPS and 70 DIAMOnD disease modules, with diseases classified into five categories using Disease Ontology (Baron et al., 2024) for disease typing. The four categories, excluding cancer, were selected based on their potential connection to proposed mechanisms of ME/CFS. The similarity measurement is based on the Jaccard index of overlapping protein interactions. (B) The network representation of the SIPS disease modules and 70 DIAMOnD disease modules, with edges colored according to the category that used the edge most frequently.

Our initial analyses offer valuable insights into the genetic makeup of ME/CFS. However, to solidify these findings, it is essential to cross-reference them with data from other ME/CFS cohorts. Thus, we incorporated data from two additional ME/CFS cohorts to enhance the SIPS disease module. The gene sets for these two cohorts were sourced from two distinct research studies:

1. The 23andMe Cohort: Based on a study published in Frontiers in Pediatrics (Perez et al., 2019a), we selected genes containing the 50 most commonly observed SNPs from gene regions in their ME/CFS cohort.
2. The UK-Nor Cohort: Using findings from a study published in Brain, Behavior, and Immunity (Hajdarevic et al., 2022), we focused on the genes proximate to their top 50 candidate variants (as detailed in the Method section).

Notably, the initial gene sets from these studies had little overlap with the SIPS seed gene set. However, as the initial gene sets expanded into disease modules, the SIPS disease module overlapped with other ME/CFS disease modules by approximately 40% (Table S3). This degree of overlap is unlikely to occur when overlapping the SIPS disease module with disease modules from random initial seeds (Figure S1).

Subsequently, we created a network representation for these three disease modules. As a reference point, we also constructed a disease module from a depressive disorder cohort (Howard et al., 2019). The disease modules from the other two ME/CFS cohorts appear to intertwine with the SIPS disease module (Figure 4A). In contrast, the depression disease module shows minimal overlap with the ME/CFS disease modules (Figure 4A). These observations suggest the possibility of shared genetic factors contributing to ME/CFS across different cohorts, and the SIPS disease module may represent a core set of genes implicated in the pathogenesis of ME/CFS.

**Figure 4.**
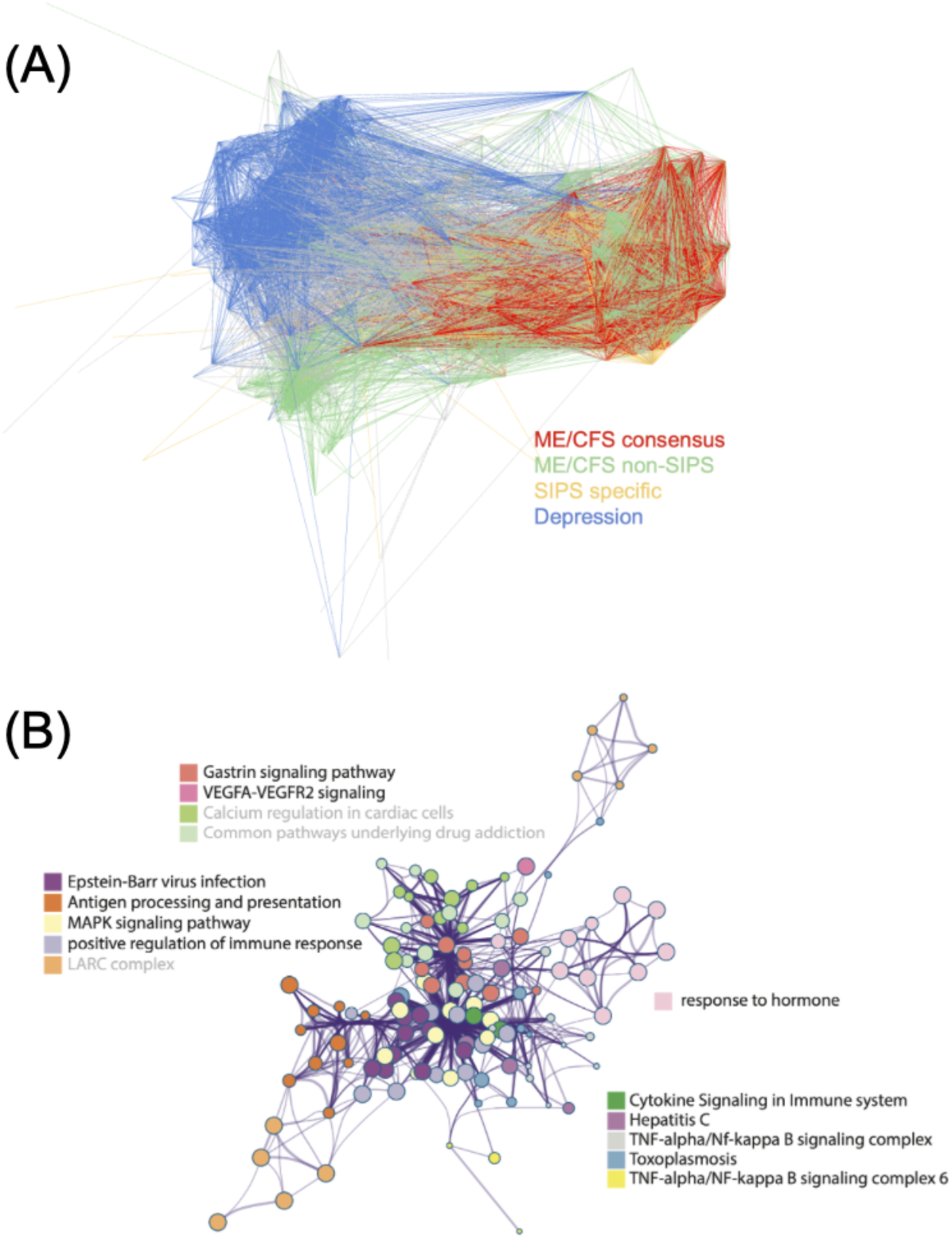
Characterization of the SIPS disease module. (A) The network representation of ME/CFS disease modules, with the depression disease module added for location reference. (B) The largest intra-cluster network component inferred by Metascape from the multi-gene set analysis, with terms not supported by all the ME/CFS disease modules being grayed out.

This finding is particularly significant in addressing the long-standing stigma surrounding ME/CFS, which has often been misdiagnosed as merely a form of depression. Such misdiagnoses have led to inadequate treatment and lack of proper medical support for ME/CFS patients. The clear distinction between the genetic underpinnings of ME/CFS and depressive disorders underscores the necessity of recognizing ME/CFS as a distinct neuroimmune condition. By elucidating these differences, our study advocates for more accurate diagnosis and targeted therapeutic approaches for ME/CFS, moving away from the erroneous conflation with depression.

To uncover shared biological components among ME/CFS disease modules, we conducted a multi-gene set analysis using Metascape (Zhou et al., 2019). The results indicated that several pathways, including those related to COVID-19, Epstein-Barr virus (EBV) infection, antigen processing and presentation, and the MAPK signaling pathway, are likely associated with ME/CFS. The top 10 biological pathway clusters linked to ME/CFS disease modules are presented in Table 1. Given the well-documented association of ME/CFS with post-viral syndromes and the increasing similarities observed between Long COVID and ME/CFS, these findings are particularly noteworthy (Komaroff and Lipkin, 2023; Ryabkova et al., 2022; Spudich and Nath, 2022). Viral infections, including EBV, have been suggested as potential triggers for ME/CFS (Deumer et al., 2021). Notably, signal impairments in the MAPK pathway in natural killer (NK) cells have been previously reported in ME/CFS patients (Chacko et al., 2016; Huth et al., 2016). Other identified pathways related to various neurodegenerative disorders and thyroid hormone signaling have also been previously associated with ME/CFS (Cortes Rivera et al., 2019; Marshall-Gradisnik and Eaton-Fitch, 2022). Interestingly, while not within the top 10 pathway clusters, the cortisol synthesis and secretion pathway was also enriched (Figure S2). This is particularly noteworthy as alterations in diurnal salivary cortisol rhythm were observed in the SIPS patients in our previous study (Chang et al., 2021).

**Table 1.**
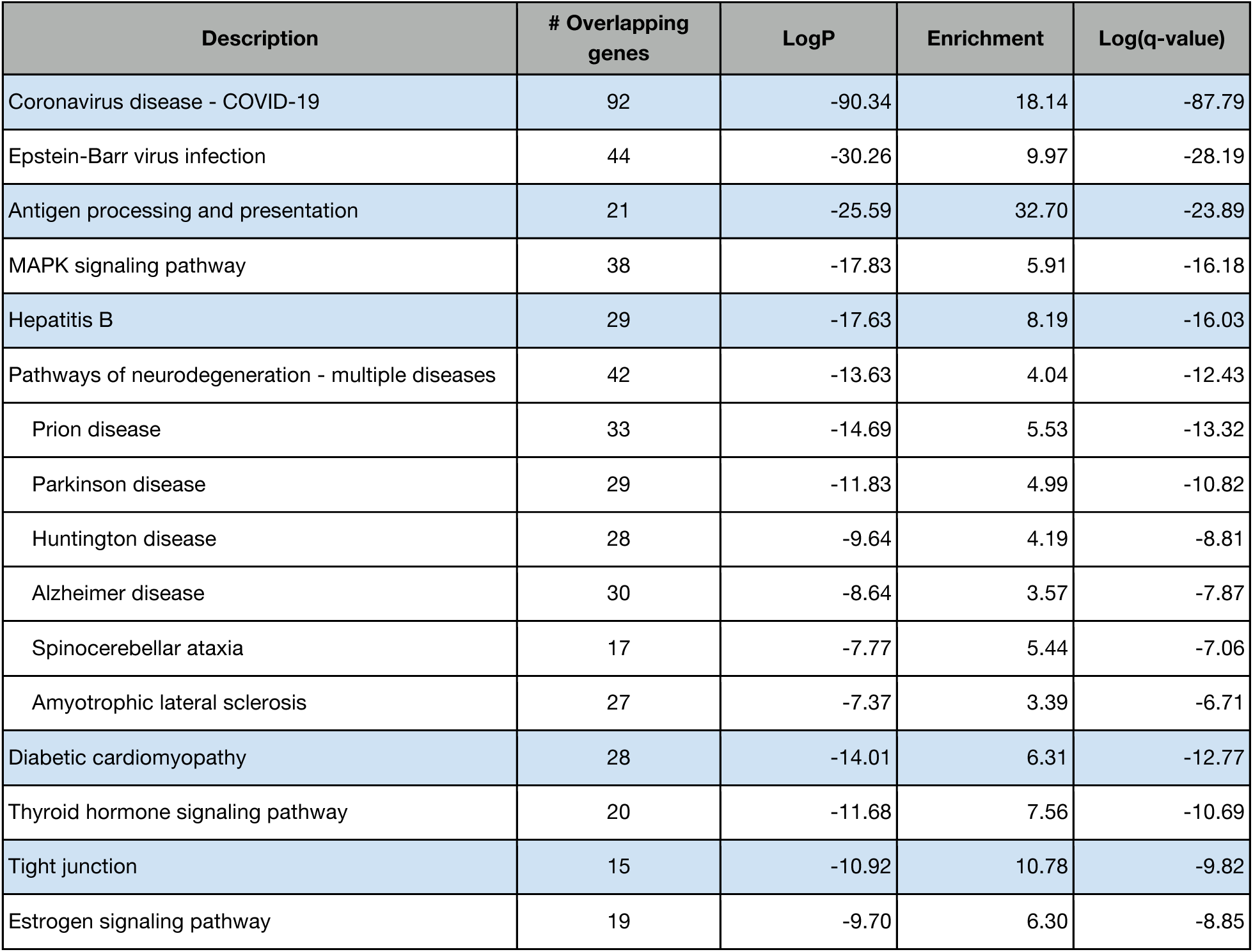
Top 10 biological pathway clusters linked to ME/CFS disease modules.

| Description | # Overlapping genes | LogP | Enrichment | Log(q-value) |
| --- | --- | --- | --- | --- |
| Coronavirus disease - COVID-19 | 92 | -90.34 | 18.14 | -87.79 |
| Epstein-Barr virus infection | 44 | -30.26 | 9.97 | -28.19 |
| Antigen processing and presentation | 21 | -25.59 | 32.70 | -23.89 |
| MAPK signaling pathway | 38 | -17.83 | 5.91 | -16.18 |
| Hepatitis B | 29 | -17.63 | 8.19 | -16.03 |
| Pathways of neurodegeneration - multiple diseases | 42 | -13.63 | 4.04 | -12.43 |
| Prion disease | 33 | -14.69 | 5.53 | -13.32 |
| Parkinson disease | 29 | -11.83 | 4.99 | -10.82 |
| Huntington disease | 28 | -9.64 | 4.19 | -8.81 |
| Alzheimer disease | 30 | -8.64 | 3.57 | -7.87 |
| Spinocerebellar ataxia | 17 | -7.77 | 5.44 | -7.06 |
| Amyotrophic lateral sclerosis | 27 | -7.37 | 3.39 | -6.71 |
| Diabetic cardiomyopathy | 28 | -14.01 | 6.31 | -12.77 |
| Thyroid hormone signaling pathway | 20 | -11.68 | 7.56 | -10.69 |
| Tight junction | 15 | -10.92 | 10.78 | -9.82 |
| Estrogen signaling pathway | 19 | -9.70 | 6.30 | -8.85 |

On the other hand, the largest intra-cluster network component identified in the multi-gene set analysis was associated with immune response pathways related to infections, such as EBV infection, cytokine signaling, positive regulation of immune response, TNF-alpha/NF-kappa B signaling, and antigen processing and presentation (Figure 4B). The network primarily focused on EBV infection and cytokine signaling, which are known to be connected to ME/CFS (Ariza, 2021; Broderick et al., 2010; Maes et al., 2022). These findings further emphasize the potential role of immune dysregulation in the pathogenesis of this disease.

## 3 Methods

### 3.1 Whole genome sequencing (WGS) data generation

WGS was performed for the 20 SIPS ME/CFS patients. Blood samples were obtained from these patients for genomic DNA extraction. The extracted DNA samples were then sequenced on an Illumina NextSeq 500 platform using a 150-bp paired-end sequencing kit.

### 3.2 Read mapping and variant calling

For each sample, 150-bp paired-end reads were aligned to the human reference genome hg19 using Isaac Genome Alignment Software (Isaac-04.16.09.24). Variant calling was performed using Strelka2 Germline Variant Caller (2.7.1), Manta Structural Variant Caller (1.0.2), and Canvas CNV Caller (1.19.1).

### 3.3 Variant filtering

High-confidence variants were derived by applying a series of filters as follows:

1. Variants had to be supported by reads from both strands.
2. Conflicting variants (i.e., multiple types of variants were called at the same position) were excluded.
3. Variants located within repetitive regions were excluded.
4. Variants in the proximity of miscalls were removed to eliminate sequencing errors in specific regions.
5. To reduce miscalls resulting from in-vitro contaminations or other systematic errors, a reasonable variant must have at least one homozygous sample in which both alleles are consistent with the reference.

In total, 2,798,019 high-confidence variants passed these five filters and were subsequently used in the following variant analysis.

### 3.4 Variant analysis

The high-confidence variants were uploaded to QIAGEN’s Ingenuity Variant Analysis (IVA) platform to identify ME/CFS-associated variants. The variant frequencies were compared with background population frequency data from the Exome Aggregation Consortium (ExAC), 1000 Genomes Project, and the Genome Aggregation Database (gnomAD).

Using the ACMG criteria for the interpretation of sequence variants, we identified a total of 103 variants as *Pathogenic* or *Likely Pathogenic*. To rule out sporadic findings from variants not yet included in these background databases, we further compared the frequencies of these variants with the Kaviar database, the aforementioned background population data, and the Allele Frequency Community (AFC; http://www.allelefrequencycommunity.com), as well as the WGS data of 20 randomly selected individuals from the Harvard Personal Genome Project (PGP). Seven of the 103 variants were removed because they had an odds ratio of less than 5 in any of these comparisons. The resulting 96 variants (103-7) were aggregated to 93 genes, with 9 genes identified that affect at least 2 (or 10%) out of the 20 of the patients.

### 3.5 Network analysis

The disease module represents a network that expands from an initial set of genes. In this study, we collected three distinct gene sets. The SIPS gene set served as our primary gene set, while the other two sets were employed for refining or validating our disease module.

1. The SIPS Gene Set: Derived from the SIPS data, this set was constructed from the 96 identified variants, encompassing 93 genes that house these variants.
2. 23andMe Gene Set: This set was retrieved from the work of Perez et al. (Perez et al., 2019b), who reported 46 genes associated with ME/CFS. These genes were extracted from the top 50 SNPs frequently reported in their ME/CFS cohort, each with C-scores exceeding 10. The 23andMe chip versions 4 (∼570 k SNPs; prior to August 2017) and version 5 (∼640 k SNPs; after August 2017) were used.
3. UK-Nor Gene Set: Based on Hajdarevic et al.’s study (Hajdarevic et al., 2022), this set was derived from the top 50 variants reported in their Supplementary Table 7. Specifically, these variants were either identified in the overlapping segments of two ME/CFS cohorts or exhibited the lowest p-values in their GWAS analysis. Genes situated within a 500K base pair radius of these variants’ linkage disequilibrium (LD) block (with r^2^ ≥ 0.5, as determined by the LDproxy function in LDLink) were chosen to represent this gene set.

## 4 Discussion

ME/CFS remains a poorly understood disease, despite its significant impact on patients’ quality of life. Our study, focusing on severely ill ME/CFS patients, provides valuable insights into potential genetic factors and pathways that may contribute to the development of this disease.

Using a network-based approach, we identified several genes and pathways potentially involved in the pathogenesis of ME/CFS, intersecting with the SIPS disease module. This module represents a network implicated in ME/CFS pathogenesis and overlaps with pathways associated with neurological diseases, which is particularly noteworthy given the prevalence of cognitive abnormalities in severely ill ME/CFS patients. Moreover, the recurring DNA variants found in genes linked to cognitive and neurological functions suggest a strong genetic basis for the cognitive irregularities observed in SIPS patients. The overlap of the SIPS disease module with other disease modules related to the immune and neurological systems further supports the hypothesis that ME/CFS is a neuroimmune disorder.

Our ME/CFS disease modules exhibit a notable enrichment of pathway related to both COVID-19 and EBV infection (Table 1). Recent reports suggest that COVID-19 could potentially reactivate EBV, a virus long suspected to play a significant role in the development of ME/CFS (Gold et al., 2021). Interestingly, many symptoms associated with Long COVID are also common in ME/CFS (Komaroff and Lipkin, 2023). This overlap could potentially be linked to the reactivation of EBV in COVID-19 patients. Consequently, inflammation-induced reactivation of EBV may contribute to some of the symptoms observed in long COVID.

Our analysis also revealed a connection between ME/CFS and estrogen signaling pathways (Table 1). This finding aligns with Perez et al.’s study conducted on the 23andMe cohort, which proposed that certain identified SNPs might participate in hormone-related pathways, including estrogen signaling (Perez et al., 2019a). Given that estrogen is known to regulate innate immune cells and signaling pathways, its potential influence on the pathophysiology of ME/CFS cannot be overlooked (Thomas et al., 2022). This could provide insights into the observed predominance of ME/CFS in females.

It is noteworthy that among the original 103 variants identified in the severely ill patients, the most prevalent is rs35767802 on the *ADAMTSL2* gene. Although this variant has >12% allele frequency according to AFC population frequency, it was present in at least one allele in 45% (9/20) of the severely affected patients. The *ADAMTSL2* gene is part of the ADAMTS (A Disintegrin and Metalloproteinase with Thrombospondin motifs) family, which plays a crucial role in the degradation of the extracellular matrix and has been linked to various human genetic disorders (Le Goff and Cormier-Daire, 2011). Notably, *ADAMTSL2* is associated with Type VIIC Ehlers–Danlos syndrome (EDS), with a mutation in *ADAMTSL2* recently identified in EDS patients (Desai et al., 2016). There may be a comorbidity between EDS and ME/CFS, as joint hypermobility—a characteristic of EDS—is frequently observed in ME/CFS patients (Eccles et al., 2021).

In addition, several biological pathways were exclusively identified in the SIPS disease module, as listed in Table S4. These pathways could potentially provide insights into the exacerbation of ME/CFS. However, we found no established connections between these pathways and ME/CFS. Pathways lacking cross-cohort validation, such as these, carry a high degree of uncertainty and may arise from methodological limitations. Therefore, further validation is needed to confirm these connections.

Overall, this study offers novel insights into the genetic factors that may contribute to severe ME/CFS, identifying several pathways potentially associated with the condition. The discovery of the SIPS disease module and its associated pathways provides a valuable foundation for future research, potentially laying the groundwork for more effective treatments for this debilitating condition. However, these findings are preliminary and, like previous studies, have limitations that necessitate further validation. The accuracy of our findings relies on the current state of the ACMG criteria for variant annotation, which may not be comprehensive. Moreover, while the DIAMOnD algorithm is widely used, its effectiveness depends on the completeness and accuracy of the underlying human PPI network data. The potential implications of these findings for the diagnosis and treatment of severe ME/CFS also need to be explored. Future studies should aim to replicate our findings in larger cohorts and to further investigate the potential role of immune and neurological pathways in severe ME/CFS.

## Data Availability

All data produced in the present work are contained in the manuscript.

## 5 Author Contributions

WZX and LYH conceived and designed the study. WZX and RWD supervised the project. LYH, CSW, and CJC collected the datasets. LYH, CSW, and CJC performed the data analysis and conducted the statistical analysis. PL, KH, BT, JJD, BM, and CLM contributed to the interpretation of the results. CSW and LYH drafted the manuscript. CSW, LYH, and WZX revised the manuscript. All authors read and approved the final manuscript.

## 7 Acknowledgments

We would also like to express our sincere gratitude to each of the severely ill patients who participated in this study.

